# SARS-CoV-2 virus transfers to skin through contact with contaminated solids

**DOI:** 10.1101/2021.04.24.21256044

**Authors:** Saeed Behzadinasab, Alex W.H. Chin, Mohsen Hosseini, Leo L.M. Poon, William A. Ducker

**Affiliations:** Dept. of Chemical Engineering and Center for Soft Matter and Biological Physics, Virginia Tech, VA, 24061, USA; School of Public Health, LKS Faculty of Medicine, The University of Hong Kong, Hong Kong Special Administrative Region, China; HKU-Pasteur Research Pole, LKS Faculty of Medicine, The University of Hong Kong, Hong Kong, China

**Keywords:** SARS-CoV-2, transfer, skin, coronavirus, COVID-19, hand, wash

## Abstract

Transfer of SARS-CoV-2 from solids to fingers is one step in infection via contaminated solids, and the possibility of infection from this route has driven calls for increased frequency of handwashing during the COVID-19 pandemic. To analyze this route of infection, we measured the percentage of SARS-CoV-2 that was transferred from a solid to an artificial finger. A droplet of SARS-CoV-2 suspension (1 µL) was placed on a solid, and then artificial skin was briefly pressed against the solid with a light force (3 N). Transfer from a variety of solids was detected, and transfer from the non-porous solids, glass, stainless steel, and Teflon, was substantial (13-16 %) when the droplet was still wet. Transfer still occurred after the droplet evaporated, but it was smaller. We found a lower level of transfer from porous solids but did not find a significant effect of solid wettability for non-porous solids.

## 1. Introduction

In the period January 2020 to March 2021, about 125 million people contracted COVID-19, and about 2.8 million have died as a result of the illness. ^1^ The disease is caused by a virus, SARS-CoV-2 (severe acute respiratory syndrome coronavirus 2). In 2020, the US CDC stated that the primary mechanism for transmission is close contacts or inhalation of respiratory droplets.^2^ Some diseases can also be transferred via contaminated solids (fomites), and the World Health Organization states that “fomite transmission is considered a likely mode of transmission for SARS-CoV-2”. ^3^A study on hamsters showed that direct inhalation is the main mechanism, but infection via fomite transmission also occurred. ^4^ Modelling of the pandemic and disease transmission found that up to 25% of the disease transmissions during lockdown was via fomites. ^5^ Studies have shown that SARS-CoV-2 remains infective up to 7 days after a droplet is placed on some solids, ^6,7^ indicating the window of possible infection from solids may be large. This has led to widespread fear of touching communal objects and to health authorities suggesting that people increase the frequency and quality of their hand-washing. ^8^ A parallel approach is to design coatings for solids that inactivate the virus and to apply these coatings to communal objects. ^9,10^

An important question remains unanswered: does the virus actually transfer from contaminated objects to a person’s hand? The transfer of virus from the contaminated solid surface to the body is necessary for infection via fomites but has not been reported. In this article, we describe measurements of the percentage transfer of SARS-CoV-2 to skin from a variety of solids.

Following biosafety protocols, we were not able to examine transfer to the skin of living humans. We instead used Vitro-skin^®^ an artificial skin for a number of reasons. First, many of its characteristics are similar to human skin. Vitro-skin^®^ has similar wettability^11^, mechanical properties^12^, textural features, and chemistry to human skin (see Supplementary Information.) Others have found Vitro-skin^®^ to be a good model for human skin. ^12,13^ An additional advantage of using an artificial skin is that human skin varies from place to place on the body, between individuals, and depends on the state of the individual. Use of a constant skin sample enables better resolution of the variability of transfer from different inanimate objects.

We made an artificial finger from polydimethylsiloxane (PDMS) with a modulus similar to human skin and attached the Vitro-skin^®^ to the PDMS. We contaminated each solid with 1 µL of SARS-CoV-2 in suspension in buffer. 1 µL is the upper end of the size range of respiratory droplets, ^14^ much smaller droplets did not allow sufficient resolution of the viral titer.

As described below, our experiments do show that SARS-CoV-2 transfers from solids to skin, even when contact is made after the droplet has evaporated.

## 2. Materials and Methods

### 2.1. Artificial Skin

Vitro-Skin^®^ was purchased from IMS Division of Florida Skincare Testing Inc. (Florida, USA). In this manuscript “artificial skin” refers to Vitro-Skin^®^. As-received Vitro-Skin^®^ needs to be hydrated to be a good model for human skin. Hydration was achieved in a sterilized, sealed chamber that contains 15% w/w glycerin in water for 16-24 hrs. ^12^ Artificial skin was sterilized with 70% ethanol prior to the hydration step.

### 2.2. Artificial finger

PDMS was chosen as support for the skin because we were able to make a PDMS support that had about the same deformation as a human finger when subjected to a 3 N load. Sylgard 184 (base to curing agent = 10:1) and Sylgard 527 (part A to part B = 1:1) were prepared separately and the two were thoroughly mixed with a ratio of Sylgard 527:184 = 5:1. Subsequently the mixture was poured into hemicylindrical-shaped molds (plastic centrifuge tube, diameter = 15 mm) and heated at 80ºC for 9 hours to cure. The resulting PDMS had a hemicylindrical shape, and the curved side faced the test solid during the transfer (See Supplementary Fig. 5). The in vivo elastic modulus of human skin is 0.04-0.22 MPa. ^15,16^ which spans the range of 0.13 MPa measured for Sylgard 527:184 = 5:1. ^17^ The sterilized and hydrated artificial skin was attached to the PDMS hemicylinder using double-sided tape to create the artificial finger.

### 2.3. Transfer experiment

A 1 µL SARS-CoV-2 droplet (at log_10_ 7.8 TCID_50_/ml) was placed on each test solid (glass, etc.) at 22-23°C and relative humidity of 60-70%, and then the artificial skin was contacted to the test solid after waiting for either 10 sec or 30 min. After 10, the droplet evaporation was negligible, and so we describe the test solid as “wet”; after 30 min, the droplet had evaporated so the test solid was dry, which we refer to as “evaporated” or “dry”. Artificial skin was attached to the curved side of the PDMS immediately before the touch experiment. Contact between artificial skin and the test solid was for 5 sec with a mass of 300 g, i.e., with a force of ≈ 3N (See Supplementary Fig. 5C). This contact force was chosen after measuring the force that we used to press a checkout credit-card machine button. The 5 sec contact time was longer than what we use to press a credit card reader button, but was chosen to achieve a smaller percentage variation in contact time. The contact area was roughly 1.2 cm x 0.3 cm, resulting in an average pressure of about 1 atm. The flat-cylinder contact was chosen to achieve a well-defined contact. A flat–flat contact would depend critically on alignment; a sphere–flat contact was not possible with the flat sheets of artificial skin that we utilized. An additional critical aspect was that that we needed to ensure that, during contact, the droplet did not run off the edges of the test solid; that would not allow accurate or reproducible accounting of the transfer. The use of a curved skin substrate provided more space within the liquid meniscus for the droplet to remain between the solids during contact. This was confirmed using dye tests prior to the experiments (Supplementary Fig. 5C).

### 2.4. SARS-CoV-2 assay

After the finger was separated from the test solid, the artificial skin was removed from the PDMS and the skin was soaked in 200 µL of viral transport medium (Earle’s balanced salt solution, which was supplemented with 0.5%(w/v) bovine serum albumin and 0.1%(w/v) glucose, pH = 7.4) at room temperature for 30 minutes to elute the SARS-CoV-2 virus (from Hong Kong index case). Subsequently, the eluted viral suspension was assayed by 50% tissue culture infective dose (TCID_50_) assay in Vero E6 cells to determine how effective the eluent was in infecting mammalian cells.^18,19^

## 3. Results and discussion

When a droplet of SARS-CoV-2 suspension is placed on a solid, the virus can be transferred to a finger, even when the finger has only a brief (5 sec) and light (3 N) touch to the solid that one may do when pressing a button. Figure 1 shows the transfer ratio, *T*, which is the ratio of the infectivity on the finger compared to the infectivity of the original droplet:

**Figure 1.**
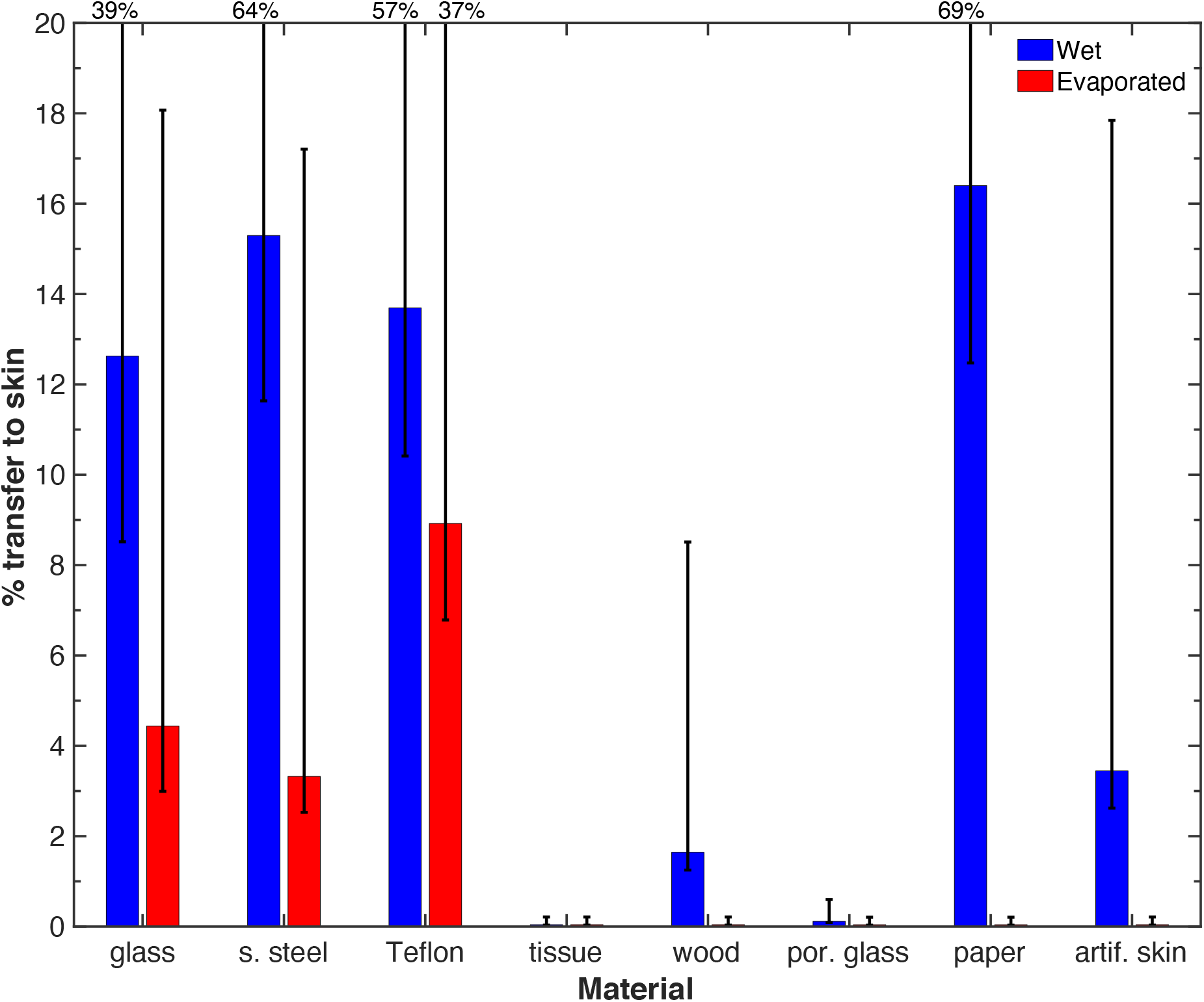
Percentage transfer to an artificial finger from various solid material during a 5 sec contact with a 3 N (300 g) load with no lateral motion (i.e., no rubbing). Error bars are the comparison intervals from ANOVA with two factors: the material and the time, which corresponds to wet or evaporated. The error bar is asymmetric because of the exponential distribution of measurement residuals. When the upper bound is off-scale, it is indicated by a number. The time to adsorb the droplet on porous surfaces was as follows: tissue and porous glass, immediate; wood, 1-2 min; and paper, 2.5 min. Note that the transfer is high for non-porous solids and is measurable when the 1 µL test drop is wet (10 sec) or has evaporated (30 min).

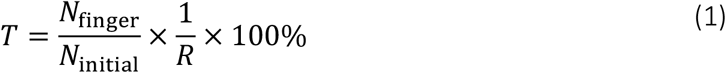

where ***N***_**finger**_ is the infectivity measured from liquid extracted from the finger after contact with the solid, as measured by TCID_50_ assay in Vero E6 (African green monkey) cells, and ***N***_**initial**_ is the infectivity measured in the droplet that was initially placed on the test solid.***R*** is the recovery rate for extracting virus from the finger. More detail is provided in Supplementary Information, S5.

We examined transfer for two conditions: (1) when it was freshly placed on the solid (after 10 sec) and (2) after it had evaporated (30 min). The transfer percentage for wet droplets on the three common non-porous solids – glass, stainless steel and Teflon – are in the range 13–16% (Fig. 1). These numbers are quite high: the amount of virus transferred to the skin after touching the solid is about 15% as great as if the droplet landed directly on the finger. When the droplet was allowed to dry, about 3–9 % was transferred. This evokes the scenario when an infected individual deposits a droplet and half an hour later a second individual who touches the dry surface could still transfer 3-9% of the original virus onto their fingers. This is not a large reduction by microbiological standards; for comparison, sanitizers and disinfectants are considered effective when they leave less than 0.1% of the germ, i.e., at least a 30–150 times greater reduction. The results reinforce the idea that hand-washing is important before touching one’s face. Note that our method of calculating the transfer includes any inactivation of the virus that occurs on the test solid prior to contact with the finger. In some cases, this could be substantial. For example, in our prior work, we showed that only 31% of the virus was recovered from glass after 30 minutes, even with no skin contact^10^. We think that the fraction of the initial dose that ends up on the finger is the relevant quantity. We note that for the transfer percentage of the dried droplets may be slightly greater than indicated, as described in Supplementary Information.

For the remaining results, we directly compare TCID_50_ values, which are shown in Fig. 2. Figure 2, Supplementary Fig. 8 and Supplementary Fig. 9 also show population marginal means. There is a significant decrease in viral transfer after the droplet evaporates (*p* <0.0001), which is more clearly shown in Supplementary Fig. 8. This is as expected, there is no liquid present to carry the virus to the finger via a meniscus.

**Figure 2.**
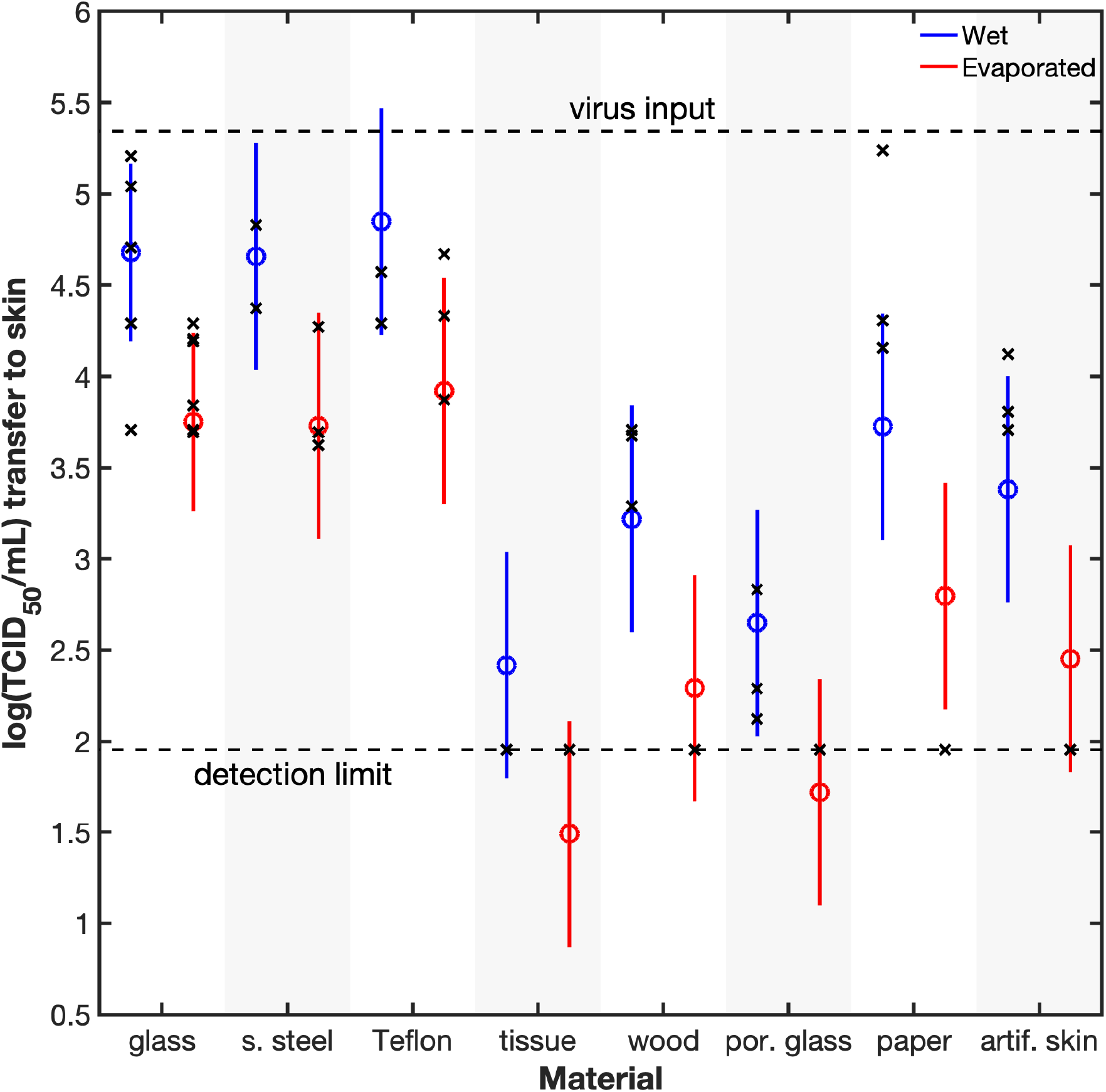
log(TCID_50_/mL) values (x) and population marginal means (o) on artificial skin after transfer from various solids. The population marginal means were calculated from Post Hoc comparison using N-way ANOVA (two factors: material and wet/evaporated) in MATLAB using the Dunn-Sidák’s approach. The bars represent the comparison interval; two groups are statistically different if their intervals do not overlap. Transfer is detectable for all solids, except tissue, wood (dry), porous glass (dry), and paper (dry), and these data points have been plotted on the detection limit of 90. Transfer is greater for wet solids than for dry and there is no significant difference among common non-porous solids.

We originally hypothesized that a more hydrophobic solid, i.e., a solid with a greater water receding angle, would transfer more virus. This was based on the idea that a greater volume of liquid would transfer from a hydrophobic solid, and therefore, more virus could be transferred in that greater volume. We examined this hypothesis by including Teflon as one of the samples.

Teflon has an unusually high receding angle (96º ± 9º) whereas clean glass has a low contact angle, 11º ± 7º (Table 1). There was no significant difference between the transfer for the two substances (see Fig. 2 and Supplementary Fig. 9). We then tested the mass of liquid that was transferred (Supplementary Fig. 7), and found that the mass transfer was also similar, which possibly explains why the viral titer did not depend on the contact angle for non-porous solids.

**Table 1.**
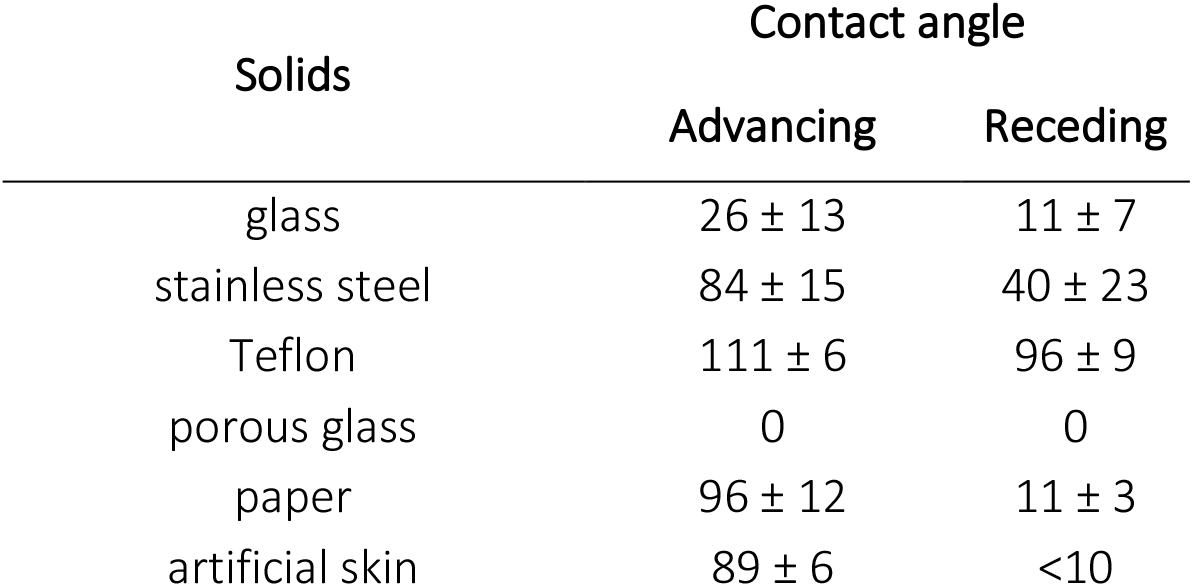
Water contact angle measurements of test solids. The figure after ± sign is the 95% confidence interval. The contact angle was measured 10 seconds after the droplet was placed on the solid.

| Solids | Contact angle |  |
| --- | --- | --- |
|  | Advancing | Receding |
| glass | 26 ± 13 | 11 ± 7 |
| stainless steel | 84 ± 15 | 40 ± 23 |
| Teflon | 111 ± 6 | 96 ± 9 |
| porous glass | 0 | 0 |
| paper | 96 ± 12 | 11 ± 3 |
| artificial skin | 89 ± 6 | <10 |

Porosity, however, does play a key role. We examined three porous objects (tissue, wood, and porous glass) that were sufficiently hydrophilic that the droplet could penetrate into the solid. Each of these porous solids had less transfer than each of the three non-porous solids (glass, stainless steel, or Teflon). This is evident for wet and dried solids considered separately (Fig. 1) and for wet and dried samples considered together (Supplementary Fig. 9). The most direct comparison is between porous and non-porous glass, where the material is the same yet the transfer is completely different. The explanation is fairly simple. For wet surfaces, the droplet is mainly within the porosity and less of the droplet is transferred to skin. For dried droplets, we expect that much of the virus is dried into the interior of the porosity and is not available on the surface for dry, solid-to-solid transfer. We did some further tests on the effect of porosity using real time quantitative polymerize chain reaction (RT-qPCR) measurements that confirm that the virus does become trapped in interior pores (see Supplementary Fig. 10). We expect that the effect of porosity would be contingent on the interior pores of the solid being wettable. Thus, we would not expect the droplet to penetrate porous Teflon, so we predict transfer would still be high from this material.

The paper material provides an interesting example of a porous material. Transfer from paper was initially (after 10 s) very high whereas transfer after the droplet evaporated (30 min) was very low. This is explained by the fact that, although paper is porous, there is an induction time for wetting: for the particular paper that we used, the paper goes from non-wetting and impermeable to wetting and permeable after about 2.5 minutes of contact with a 1 µL droplet (See Supplementary Figure 12) and the water can permeate into the solid. Our “wet” experiment was conducted after 10 s, during the impermeable period. Without access to the porosity, the SARS-CoV-2 suspension behaved like it was on a non-porous solid, the droplet was transferred to the finger and we recorded a high transfer ratio. After 2.5 min., the viral suspension penetrated into the paper, and subsequently dried into the pore space, presumably depositing the virus onto the paper fibers on the interior of the paper. The transfer experiment after 30 minutes showed a very low level of transfer because those interior surfaces were not accessible to the finger during transfer.

A further interesting aspect of porosity concerns the fate of the SARS-CoV-2 in the porosity. We also measured the TCID_50_ of liquid used to extract virus from the <u>dried solids</u> after contact with the finger (data not shown) using the same procedure as for extraction from the skin (see section 5.4.5). We were not surprised that the levels were very low for porous solids. For porous glass, tissue, and wood, TCID_50_ was below the detection limit. However, for paper, log[TCID_50_/mL] was 3.5. This indicates that SARS-CoV-2 can, to some extent, be stored in dried paper, and resuspended in an active state.

Finally, we used linear regression to make a predictive model for the transfer as a function of the wet/dry state, the porosity, and the wettability. This model performed well and is described in Supplementary Fig. 13.

## 4. Discussion

People use a huge range of contact trajectories when touching surfaces, for example, different pressures, different times of contact, and different rubbing actions. In this study we considered only brief and low-pressure contact with no rubbing or translation in contact. It is well-established that rubbing causes friction and increases wear and transfer between solids, and in particular that microbial transfer is increased significantly by rubbing.^20^ We therefore expect that transfer of SARS-CoV-2 would also be enhanced by rubbing.

Infected patients can shed droplets with high concentrations of SARS-CoV-2. ^21,22^ For example, Pan *et al*. showed a median of 7.5 × 10^5^ and a high of 10^7^ gene copies per mL in sputum. ^21^ Our research shows that, on non-porous solids, 13–16% of this virus can be transferred to skin after being deposited on a solid when the drop is still wet and 3–9% can be transferred even 30 minutes later, when the droplet is dry. This still leaves a large dose of virus on the finger. Clearly one more step is required to transfer the virus to the respiratory system: transfer from the finger to the nose or mouth. The overall transmission from solid to skin to respiratory system depends on the product of the individual transfer rates, and would be lower. The final dose transferred to the respiratory system should be compared to the infectious dose for a human, which unfortunately is not known. Studies of Syrian hamsters indicate that the infective does for these susceptible animals is only 5 infectious particles. ^23^ Clearly hand washing can reduce both the contamination of surfaces and intercept the virus before it is transferred from fingers to the respiratory system.

Infection via fingers depends also on the longevity of the virus on human skin: the virus must survive long enough on skin to be transferred. Harbourt et al. reported the stability of SARS-CoV-2 on dead porcine (pig) skin. ^24^ They found the virus remains stable for 4 days at 22±2°C, and 8 h at 37±2°C (relative humidity of 40-50%). Hirose et al. measured survival of the SARS-CoV-2 virus on a dead human skin model. ^25^ They showed the virus can survive on the skin model for up to 9 hours at 25°C and relative humidity of 45–55%. These longevity studies suggest that once SARS-CoV-2 has been transferred to the skin, there is a long period of vulnerability.

In summary, our research demonstrates substantial transfer of SARS-CoV-2 virus from a variety of solids to an artificial finger, even when the surface has dried after application of a viral suspension and for a brief, light force and no rubbing. When the droplet penetrates a porous solid, such as wood or tissue, the transfer is low.

## Data Availability

The data that supports the findings of this study are available within the Supplementary Material.

## Supplementary Material

See Supplementary Material for further information about Supplementary Materials and Methods, Characterization of Samples, Transfer Apparatus, Statistical analysis, Data Analysis, Mass Balance for Transfer Efficiency, Calculation of the transfer ratio by another method, Data tables, and Additional figures.

## Author Contributions

WD and LLMP conceived of the experiments and WD designed the transfer apparatus. WD and SB designed the artificial finger. SB contributed to design of experiment, prepared and characterized the samples unless otherwise noted. MH fabricated the porous glass samples.

Analysis of data was done by WD, SB, AWHC, MH, and LLMP. The preparation of SARS-CoV-2 and experiments containing the virus were done by AWHC. Supervision was done by WAD and LLMP. The manuscript was written by SB and WD and MH with input from all authors.

## Acknowledgments

The authors acknowledge use of Microscopy facilities within the NCFL at Virginia Tech. We thank Professor Thomas Staley for grinding the glass particles to an appropriate size, Kevin Holshouser for fabricating the transfer apparatus, and Matty Ducker for the linear regression. This work was supported by the National Science Foundation under Grant No. CBET-1902364, the Health and Medical Research Fund (COVID190116), and the National Institute of Allergy and Infectious Diseases (contract HHSN272201400006C)

## Data availability

The data that supports the findings of this study are available within the Supplementary Material.

