## Supplementary Material for "SARS-CoV-2 virus transfers to skin through contact with contaminated solids"

### S1. Supplementary Materials and Methods

#### S1.1. Materials

The following items were purchased from VWR: glass slides (25 × 75 × 1 mm), acetone (ACS grade), nitric acid (70%, ACS grade), 70% ethanol, and ethanol (200 Proof). Polydimethylsiloxane (PDMS) Sylgard 184 and 527 were obtained from Dow Corning. Tissue was purchased from Vinda (Hong Kong). Wood was part of a disposable tongue depressor purchased from Navy (Hong Kong). The test solid of paper was TRU RED™ printer paper (SKU# 13585, 75 g/m$^2$). Water was purified by Milli-Q Reference system. Glass slides were cut into 12x12 mm, rinsed with water (3 times), immersed in 70% ethanol for 15 min. Subsequently, rinsed with water (3 times), soaked in 6 M nitric acid for 20 min and rinsed with water. Fatigue-resistant 301 stainless steel (301 SS, 6”x6”, ½ hard temper, 0.015” thick) and chemical resistant PTFE (Teflon) sheet (12”x12”x1/16”) were purchased from McMaster-Carr. 301 SS was cut into 12x12 mm pieces, washed with soapy water, rinsed with water. Teflon was cut into 12x12 mm pieces, washed with a soap solution in water, rinsed with water, immersed in and rinsed with ethanol and then acetone.

#### S1.2. Methods

##### S1.2.1. Characterization

Contact angles were measured using a First Ten Angstroms FTA125. Scanning Electron Microscope (SEM, JEOL IT500 SEM) was used to characterize the morphology. The porous glass sample was sputtered with 5 nm of Au/Pt prior to SEM imaging.

The following methods were used to determine the similarity between Vitro-Skin® (artificial skin) and human skin: Attenuated Total Reflectance-Fourier Transform Infrared (ATR-FTIR, Varian 670-

FTIR), optical microscopy (Zeiss Axio Imager.M2 upright fluorescent microscope equipped with a 10 × objective).

#### S1.2.2. Fabrication of porous glass test solids

Smooth crushed glass (270-1000 grit, catalog number 64223704) was purchased from MSC Industrial Supply Company. The crushed glass was milled in U.S. Stoneware roller mill with alumina milling media at 0.5 rotations per sec. The attachment of the particles to glass slides was accomplished as follows. A suspension of 10.5 wt.% milled glass in ethanol was sonicated for five minutes, 280 μL of suspension applied on 15×15 mm pieces of glass slide, and then the samples were dried at room temperature for 30 minutes. Early-stage sintering of particles to each other and to the slide was achieved by heat treatment at 120$^{o}$C for 10 minutes, then at 320$^{o}$C for 10 minutes, and finally at 617$^{o}$C for 2 hours. Next, the furnace was switched off and samples were cooled gradually to room temperature. Finally, the porous glass test solids were cleaned in the same way as the glass slides.

#### S1.2.3. Preparation of SARS-CoV-2 culture

Our viral assay methods were described previously.[1-3] In summary, all of the surfaces were sterilized with 70% ethanol and dried in air before testing with the virus, unless otherwise noted. The SARS-CoV-2 strain was from the index case of Hong Kong. The stock virus was prepared in Vero-E6 cells cultured in Dulbecco's Modified Eagle Medium (DMEM, with 2% fetal bovine serum and 1% v/v penicillin-streptomycin) at 37°C with 5% $CO_2$.

#### S.1.2.4. SARS-CoV-2 assay

After the finger was separated from the test solid, the artificial skin was removed from the PDMS and the skin was soaked in 200 µL of viral transport medium (Earle's balanced salt solution, which was supplemented with 0.5%(w/v) bovine serum albumin and 0.1%(w/v) glucose, pH = 7.4) at room temperature for 30 minutes to elute the SARS-CoV-2 virus (from Hong Kong index case). Subsequently, the eluted viral suspension was assayed by 50% tissue culture infective dose ($TCID_{50}$) assay in Vero E6 cells to determine how effective the elutant was in infecting mammalian cells.[4,5]

Briefly, the liquid containing the virus was serially diluted in quadruplicates and infected confluent Vero E6 cells on 96-well plates. Subsequently, the infected cells were incubated at 37°C with 5% $CO_2$. On day 5 post-infection, the cells were examined for a cytopathic effect. The $TCID_{50}$/ml is the dilution that caused a cytopathic effect in 50% of treated Vero E6 cell cultures (N=4 per each dilution; Reed-Muench method[6]). Three independent tests were done at each condition. Error residuals were approximately normally distributed after a log transformation, so all statistics were calculated from the log of the titers

#### S.1.2.5. qPCR test

Viral RNA was extracted from 70 ml of VTM containing the virus using QIAamp Viral RNA Mini Kit (Qiagen) and eluted in 30 ml elution buffer. Two ml of the eluted viral RNA was subjected to quantitative RT-PCR to quantify the N gene of SARS-CoV-2 using TaqMan Fast Virus 1-step Master Mix (Thermo Fisher). The primers and probe for the assay are: 5′-

TAATCAGACAAGGAACTGATTA-3′ (Forward), 5′-CGAAGGTGTGACTTCCATG-3′ (Reverse) and 5′-GCAAATTGTGCAATTTGCGG-3′ (Probe in 5’-FAM/ZEN/3’-IBFQ format), as described in.[7]

### S2. Characterization of Samples

#### S2.1. Vitro-Skin®

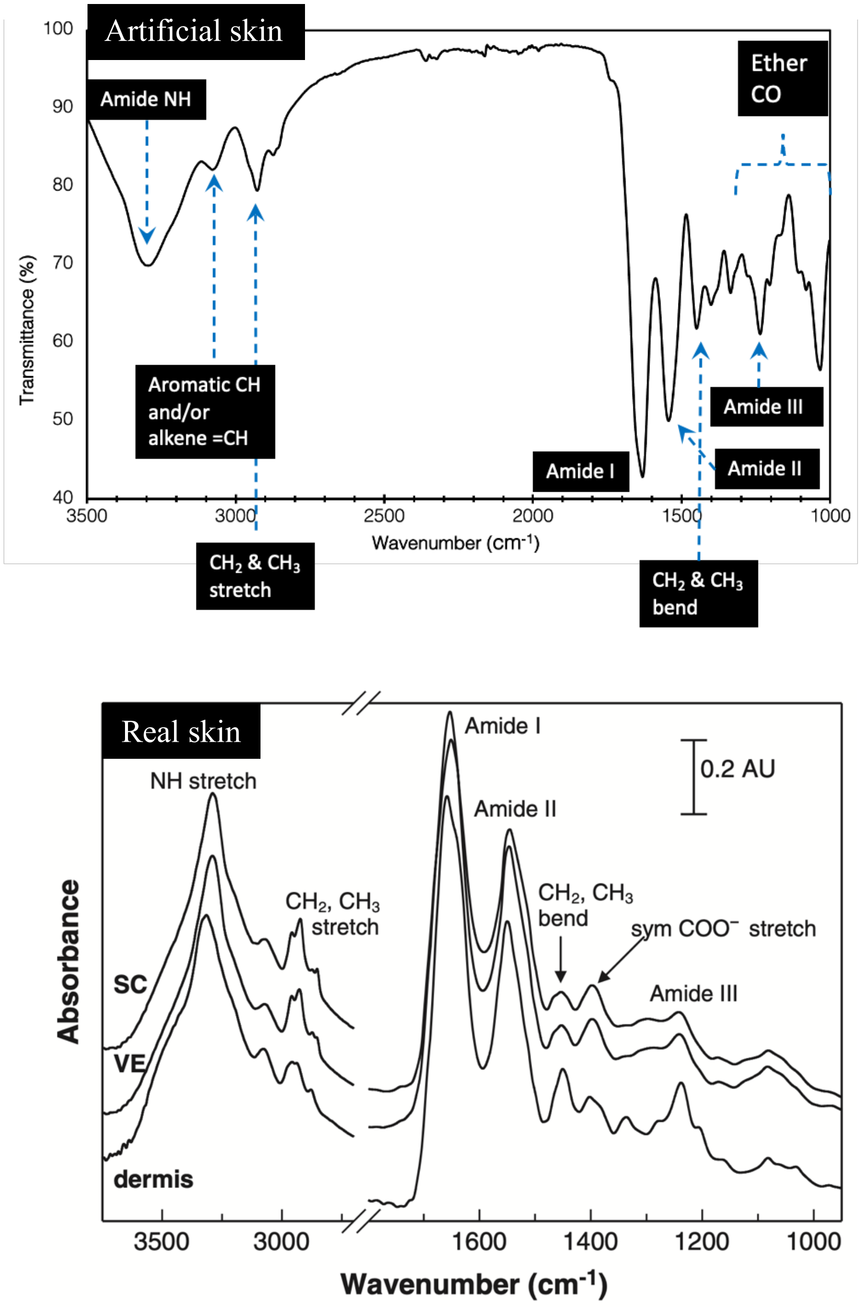

**Supplementary Figure 1. IR spectra of (top) artificial skin and (bottom) real human skin.** Note that the Vitro-Skin® data is presented as transmittance, whereas the real skin data is presented as absorbance. The artificial skin and real human skin have similar IR spectra, which is consistent with similar chemistry. (IR of real skin was reused with permission from Ref. [8], © 2012 Wiley).

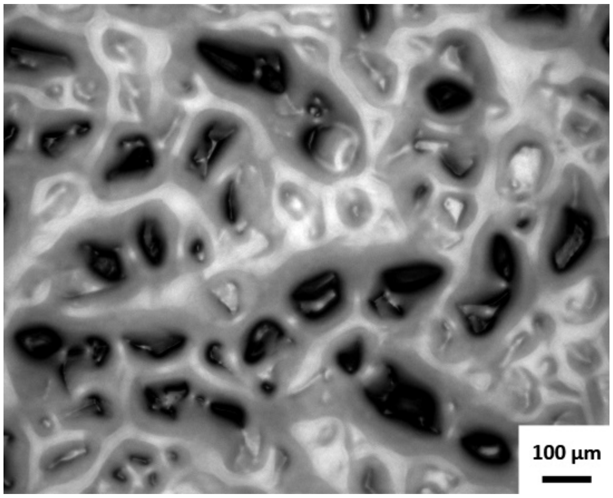

Supplementary Figure 2. Optical image of Vitro-Skin®.

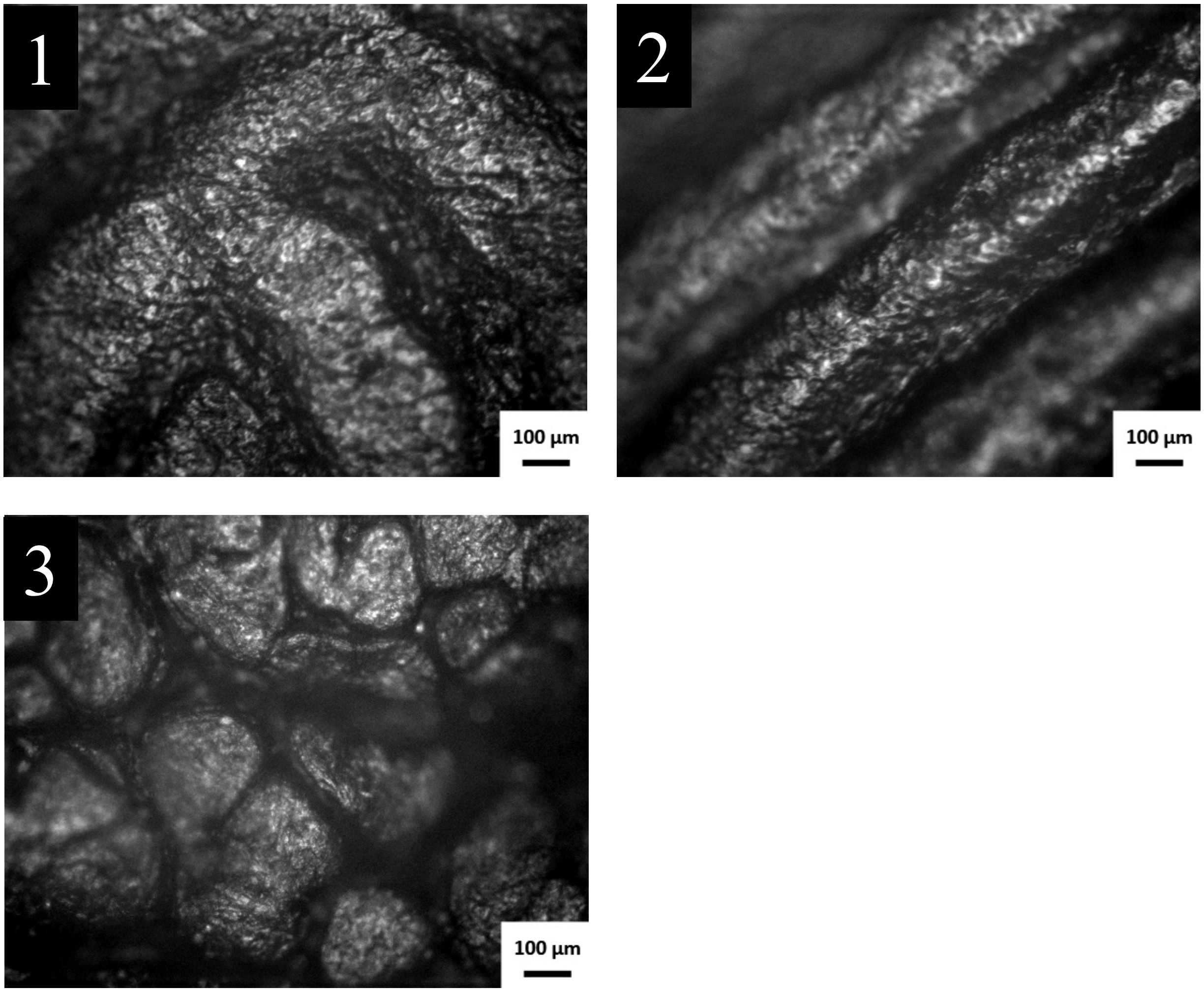

**Supplementary Figure 3. Optical images of human hand skin.** Number 1 and 2 show two regions of the finger-print region of index finger of a right-hand. Number 3 shows skin on the nail-side of the right-hand index finger. Human skin topography varies depending on the location. The subject’s hand was washed with antibacterial soap (twice) immediately before imaging. The feature size is similar on human skin and Vitro-Skin®, but Vitro-Skin® does not have fingerprints.

S2.2. Porous Glass

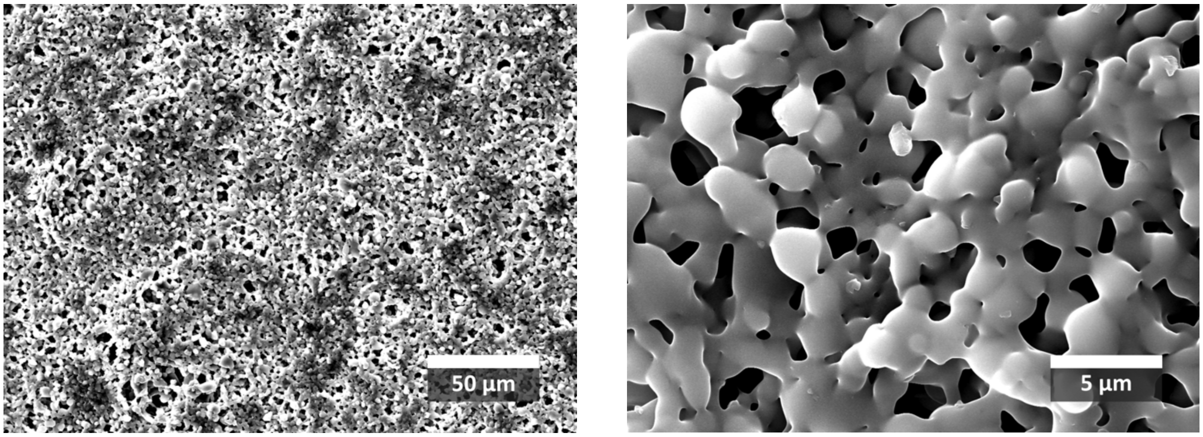

**Supplementary Figure 4. SEM images of the porous glass sample.** The images show the pore size ranges from sub-µm to above to a few µm.

#### S3. Transfer Apparatus

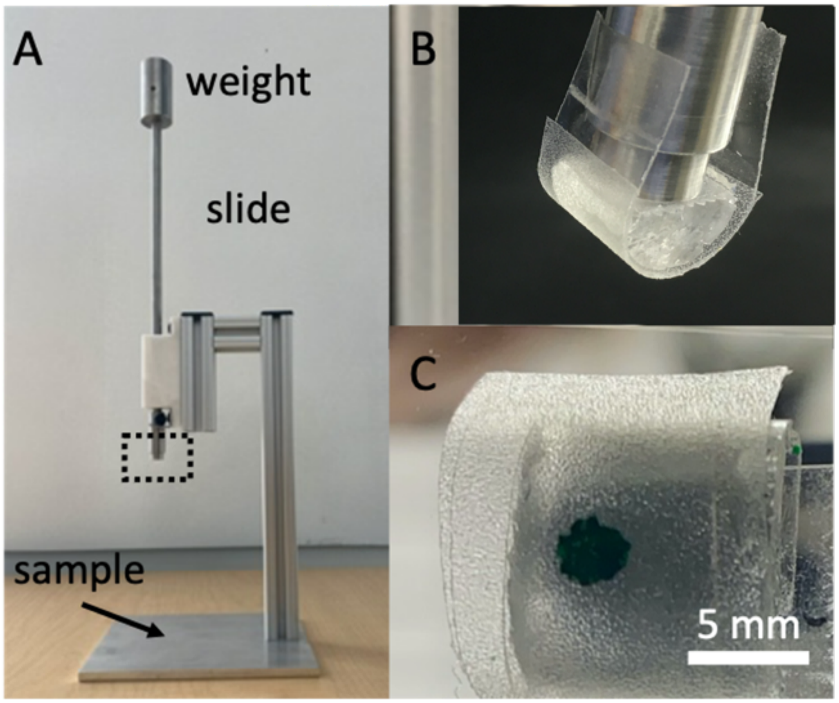

**Supplementary Figure 5. Photographs of apparatus.** (A) Device for applying 3 N constant force. Square box shows area of detail in (B). (B) higher magnification view of the imitation finger showing semicircular cross-section. (C) The finger in contact with a glass slide. The slide was moved down so that the artificial finger contacted the sample with a known force of 3 N. For this photograph, the droplet was dyed green for easier visibility. The PDMS cylinder is flattened by the load.

#### S4. Statistical analysis

We and others find that the errors in log ($TCID_{50}$) are distributed normally, and therefore all statistics were obtained after a log transformation of the raw $TCID_{50}$ data. Data for Fig. 2, Supplementary Fig. 8 and Supplementary Fig. 9 was analyzed using N-way ANOVA in MATLAB with two factors, material (glass, stainless steel, etc.) and wet/evaporated. The population marginal means were estimated using Post Hoc multiple comparison (Dunn-Sidák's approach). Additionally, we did a regression analysis with three factors – porosity (categorical), wettability (numerical from contact angle) and wet/evaporated. $p < 0.001$ for coefficients for porosity and wet/dry

### S5. Data Analysis

As described in the manuscript, we defined Transfer Efficiency as:

$$T = \frac{N_{\text{skin}}}{N_{\text{initial}}} \times \frac{1}{R} \times 100\% \qquad \text{(Supplementary Eq. 1)}$$

where $N_{\text{skin}}$ is the $TCID_{50}$ measured on skin after contact with the solid and $N_{\text{initial}}$ is the $TCID_{50}$ measured in the droplet that was initially placed on the solid. The $TCID_{50}$ technique is not a measure of viral numbers but a measure of infectivity. The exact relationship between $TCID_{50}$ and number of virions is unknown, but if we assume that it is linear, then the proportionality cancels in S1 and the Transfer ratio represents the fraction of virions that are transferred. When comparing $N_{\text{skin}}$ to $N_{\text{initial}}$ it is important to note that $N_{\text{skin}}$depends on our ability to recover the virus from a solid, whereas $N_{\text{initial}}$ does not, so we need to account for fraction of virus recovered from the skin, $R_{\text{skin}}$, which was calculated as follows (see Supplementary Fig. 6). This was determined in a separate experiment by placing a viral suspension of known $TCID_{50}$ directly on the artificial finger (i.e., no contact with a test solid) and then eluting the droplet in the same way as was done for skin after each transfer experiment. $R_{\text{skin}}$was calculated from:

$$R_{\text{skin}} = \frac{N_{\text{skin}}}{N_{\text{initial}}} \qquad \text{(Supplementary Eq. 2)}$$

where $N_{\text{skin}}$ is the $TCID_{50}$ for the liquid eluted from the skin, and $N_{\text{initial}}$ is the $TCID_{50}$ from the original droplet that was placed on the skin. In principle, $N_{\text{initial}}$ from Supplementary Eq. 1 and Supplementary Eq. 2 could be measured at the same time, but in practice they were measured at different times and therefore are slightly different. Note that the $N$ values in Supplementary Eq. 1 and Supplementary Eq. 2 are the averages of 3 independent $TCID_{50}$measurements, and because the residuals are distributed normally after a log transformation are calculated from:

$$N = \text{antilog}\left[\text{average}\left(\log(\text{TCID}_{50})\right)\right] \qquad \text{(Supplementary Eq. 3)}$$

The value of $R_{\text{skin}}$ was 0.99 and $\text{TCID}_{50}$ of virus recovered from artificial skin after is not significantly different than from the droplet was placed on the skin. We have not devised a method for determining $R_{\text{skin}}$ for a solid when dried virus is transferred so we have used the value of $R_{\text{skin}}$ for the transfer from a wet solid. We expect that the extraction will be more difficult for dried virus so we consider the reported transfer for virus from various dried solids to be lower bounds.

We and others find that the errors in log ($\text{TCID}_{50}$) are distributed normally, and therefore all statistics were obtained after a log transformation of the raw $\text{TCID}_{50}$ data, i.e.:

$$N = \text{antilog}\left[\text{average}\left(\log(\text{TCID}_{50})\right)\right] \qquad \text{(Supplementary Eq. 3)}$$

### S6. Mass Balance for Transfer Efficiency

To validate our approach for Supplementary Eq. 1, we examined the mass balance for the virus for transfer from two solids, the glass and the stainless steel. For these two materials, we have $R_{\text{fraction}}$ from prior work[3]. If no virus is lost and our assumptions for Supplementary Eq. 1 were correct, then the sum of the amount recovered from the donor surface and from the skin should equal the amount originally added to the donor. To utilize the $\text{TCID}_{50}$ data we must make the additional assumption that the $\text{TCID}_{50}$ values are additive, which rests on the unproved idea that the dose response of the Vero E6 cells to SARS-CoV-2 is linear. Making this assumption:

$$\mathrm{TCID_{50}(final)} = \frac{\mathrm{TCID_{50}(donor)}}{R_{\mathrm{donor}}} + \frac{\mathrm{TCID_{50}(recipient)}}{R_{\mathrm{recipient}}} \quad \text{(Supplementary Eq. 4)}$$

Supplementary Eq. 4 again assumes that the $\mathrm{TCID_{50}}$ values are additive. Supplementary Figure 6 is a schematic of the mass balance experiment.

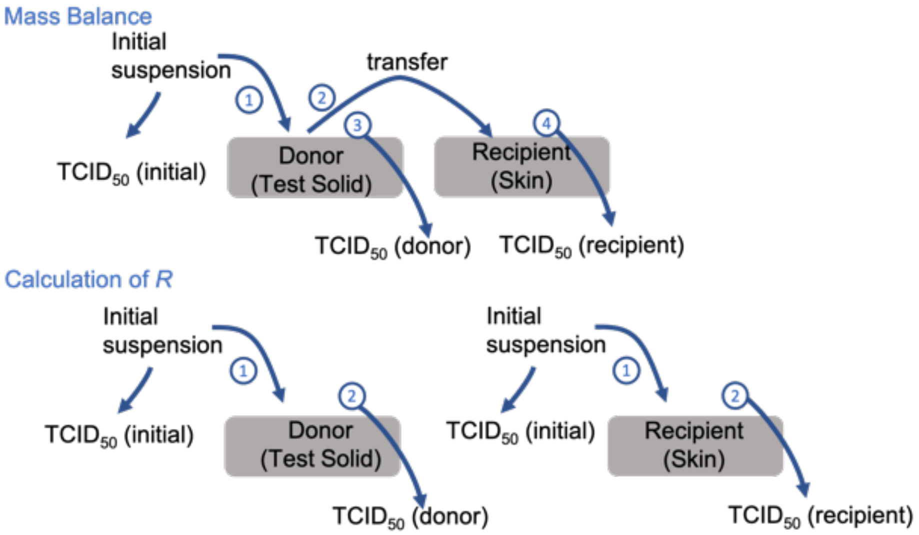

**Supplementary Figure 6. schematic of the mass balance experiment**

Supplementary Table 2 shows relevant data for the calculation. We compare the $\mathrm{TCID_{50}}$ (final) from Supplementary Eq. 4, to the $\mathrm{TCID_{50}}$ of the suspension droplet that we placed on the surface (initial). For wet glass we obtain 44% in the first measurement and 41% in the second experiment. For dry glass, we account for 109% in the first experiment and 170% in the second experiment. For wet stainless steel, we account for 33%. We do not have data to calculate mass balance for the other combinations. Possible reasons for the lack of mass balance are that the dose-response is highly non-linear and it is not valid to perform the sum in Supplementary Eq. 4. Also, there is a large uncertainty in R values. We note that we do not use Supplementary Eq. 4 in our data analysis.

### S7. Calculation of the transfer ratio by another method.

Another potential method for calculating transfer ratio is from: $\frac{\frac{\mathrm{TCID}_{50}(\mathrm{recipient})}{R_{\mathrm{recipient}}}}{\left(\frac{\mathrm{TCID}_{50}(\mathrm{recipient})}{R_{\mathrm{recipient}}}+\frac{\mathrm{TCID}_{50}(\mathrm{donor})}{R_{\mathrm{donor}}}\right)}$.

We reject this method because:

(a) The TCID values are not necessarily additive, making the calculation in the denominator problematic.

(b) There are very large uncertainties in calculating R and therefore in correctly weighting the various terms. The mass balance calculation in Section S6 demonstrates this.

(c) There is large variation in the measured values of transfer ratios for repeat measurements.

### S8. Data tables

| | | TCID50/ml | | | | log (TCID50/ml) | | |
|---|---|---|---|---|---|---|---|---|
| Nil (virus input control) | | 419371.2 | 160669 | 160669 | | 5.622599 | 5.205932 | 5.205932 |
| Material | | | | | | | | |
| Glass (wet) | | 5080.798 | 5080.798 | 160669 | | 3.705932 | 3.705932 | 5.205932 |
| Glass (dry) | | 6927.034 | 5080.798 | 15587.41 | | 3.840547 | 3.705932 | 4.192774 |
| Stainless steel (wet) | | 23582.98 | 23582.98 | 67753.53 | | 4.372599 | 4.372599 | 4.830932 |
| Stainless steel (dry) | | 18694.83 | 4193.712 | 4929.173 | | 4.271721 | 3.622599 | 3.692774 |
| Teflon (wet) | | 19465.49 | 37266.33 | 37266.33 | | 4.289265 | 4.571317 | 4.571317 |
| Teflon (dry) | | 7457.592 | 46796.96 | 21425.55 | | 3.872599 | 4.670218 | 4.330932 |
| Artificial skin (wet) | | 13261.68 | 5080.798 | 6396.346 | | 4.122599 | 3.705932 | 3.805932 |
| Artificial skin (dry) | | < | < | < | | < | < | < |
| Tissue (wet) | | < | < | < | | < | < | < |
| Tissue (dry) | | < | < | < | | < | < | < |
| Wood (wet) | | 4728.049 | 1946.549 | 5080.798 | | 3.674682 | 3.289265 | 3.705932 |
| Wood (wet) | | < | < | < | | < | < | < |
| Nil (virus input control) | | 160669 | 194654.9 | 372663.3 | | 5.205932 | 5.289265 | 5.571317 |
| Material | | | | | | | | |
| Glass (wet) | | 19465.49 | 109462.5 | 50807.98 | | 4.289265 | 5.039265 | 4.705932 |
| Glass (dry) | | 4929.173 | 19465.49 | 16066.9 | | 3.692774 | 4.289265 | 4.205932 |
| Porous glass (wet) | | 194.6549 | 677.5353 | 132.6168 | | 2.289265 | 2.830932 | 2.122599 |
| Porous glass (dry) | | < | < | < | | < | < | < |
| Paper (wet) | | 172656.1 | 14319.64 | 20227.02 | | 5.237182 | 4.155932 | 4.305932 |
| Paper (dry) | | < | < | < | | < | < | < |

**Supplementary Table 1. Measured $TCID_{50}$/mL values of the transferred virus to the skin.** The “<” sign represents the titer measured was below the detection limit of the $TCID_{50}$ assay, which is at 90 $TCID_{50}$/ml. We used the value of 90 during statistical analysis, so this is an upper bound.

| | | TCID50/ml | | |
|---|---|---|---|---|
| Nil (virus input control) | | 419371.2 | 160669 | 160669 |
| Material | | | | |
| Glass (wet) | | 5080.798 | 5080.798 | 160669 |
| Glass (dry) | | 6927.034 | 5080.798 | 15587.41 |
| Stainless steel (wet) | | 23582.98 | 23582.98 | 67753.53 |
| Stainless steel (dry) | | 18694.83 | 4193.712 | 4929.173 |
| Nil (virus input control) | | 160669 | 194654.9 | 372663.3 |
| Material | | | | |
| Glass (wet) | | 19465.49 | 109462.5 | 50807.98 |
| Glass (dry) | | 4929.173 | 19465.49 | 16066.9 |

Supplementary Table 2. Values used for calculation of mass balance (see section S5.). $R_{\mathbf{donor}}$ for glass (wet) and glass (dry) are 0.905 and 0.307, respectively. That for stainless steel (wet) is 0.747. $R_{\mathbf{recipient}}$ for skin (wet) and skin (dry) are 0.991 and 0.041, respectively.
The calculation were done as follow: the R's were applied to the raw TCID50 values, the values were calculated according to Supplementary Eq. 3, after which Supplementary Eq. 4 was used to do the mass balance.

S8. Additional figures

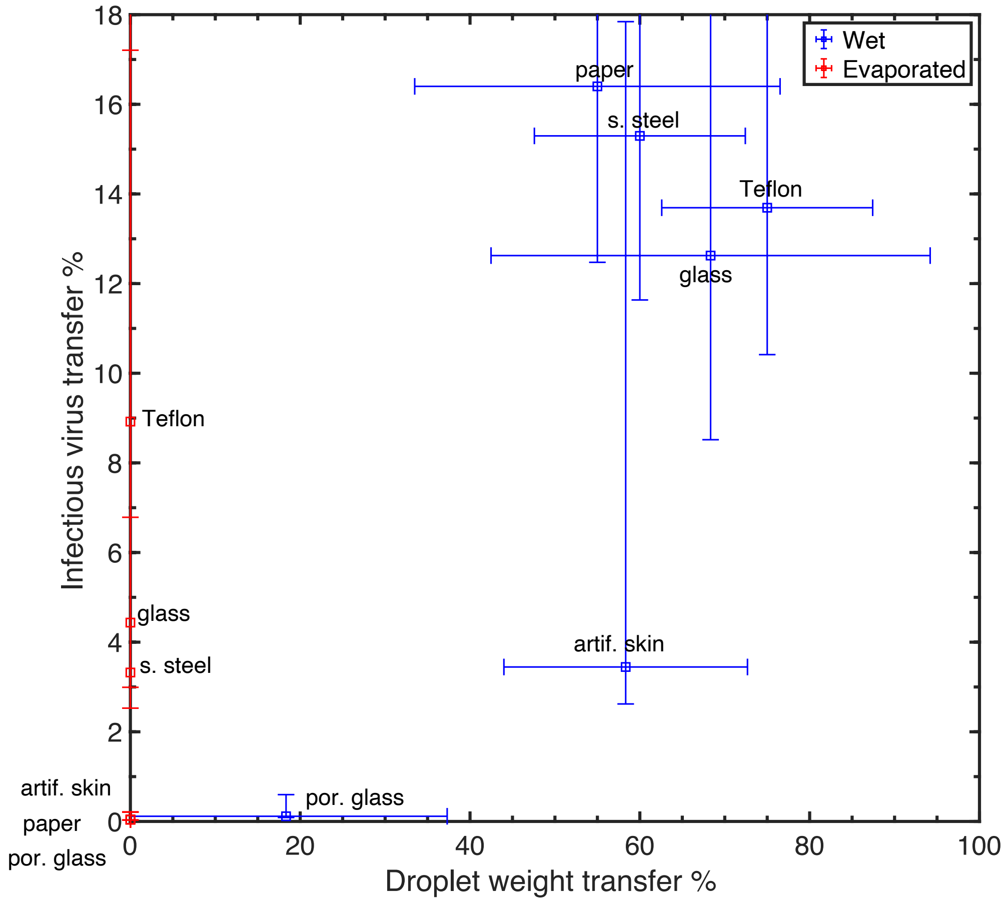

**Supplementary Figure 7. Percentage of droplet mass transferred to a finger from artificial skin after the skin was contacted to various solids.** The transfer experiment was repeated but, this time the test droplet was 2 mg of DMEM without any virus and the measurement was the mass instead of the $TCID_{50}$. This was to test whether the differences in transfer were entirely due to transfer of the liquid. Note that there is not a significant difference between the results for wet Teflon and wet glass, in spite of the large difference in receding contact angle. The very low percentage mass transferred in the dry experiments occurs because the droplet has evaporated and the mass of dissolved solid and suspended virus is minute. Clearly the wet samples transfer more virus and more water, the Pearson correlation coefficient between mass and transfer is $R^2$= 0.57. Considering only the wet experiments, the amount of water transferred is still not a great predictor of the virus transfer ($R^2$= 0.50), and the correlation is even lower if the porous glass is excluded.

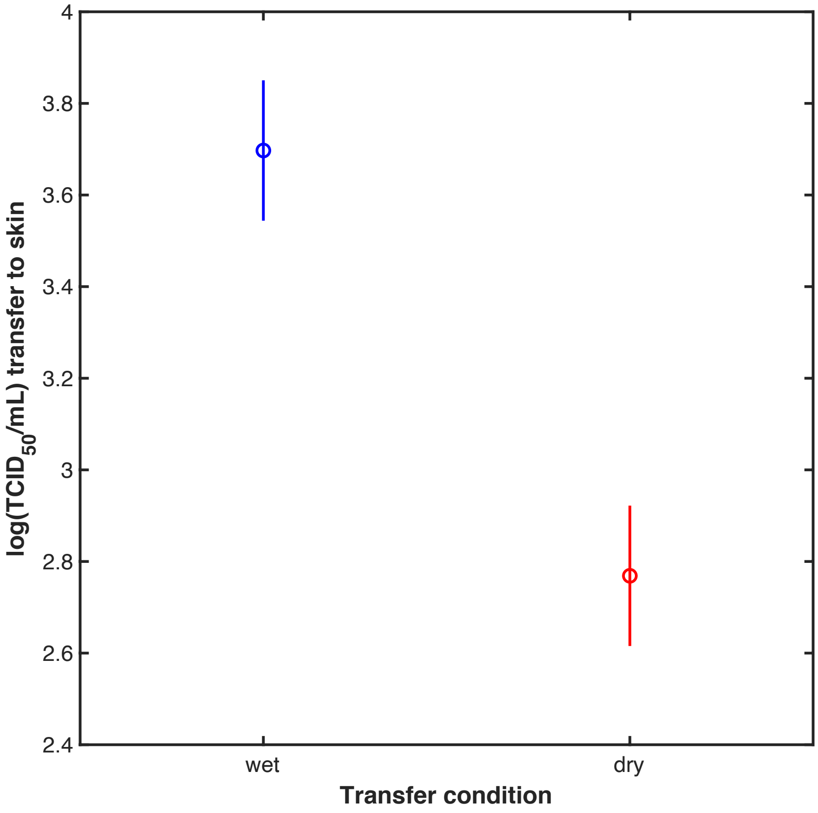

**Supplementary Figure 8. Post Hoc comparison of log($TCID_{50}$/mL) for skin after contact when the droplet is wet and when it has evaporated (dry).** More virus is transferred to skin when the droplet of virus suspension is wet.

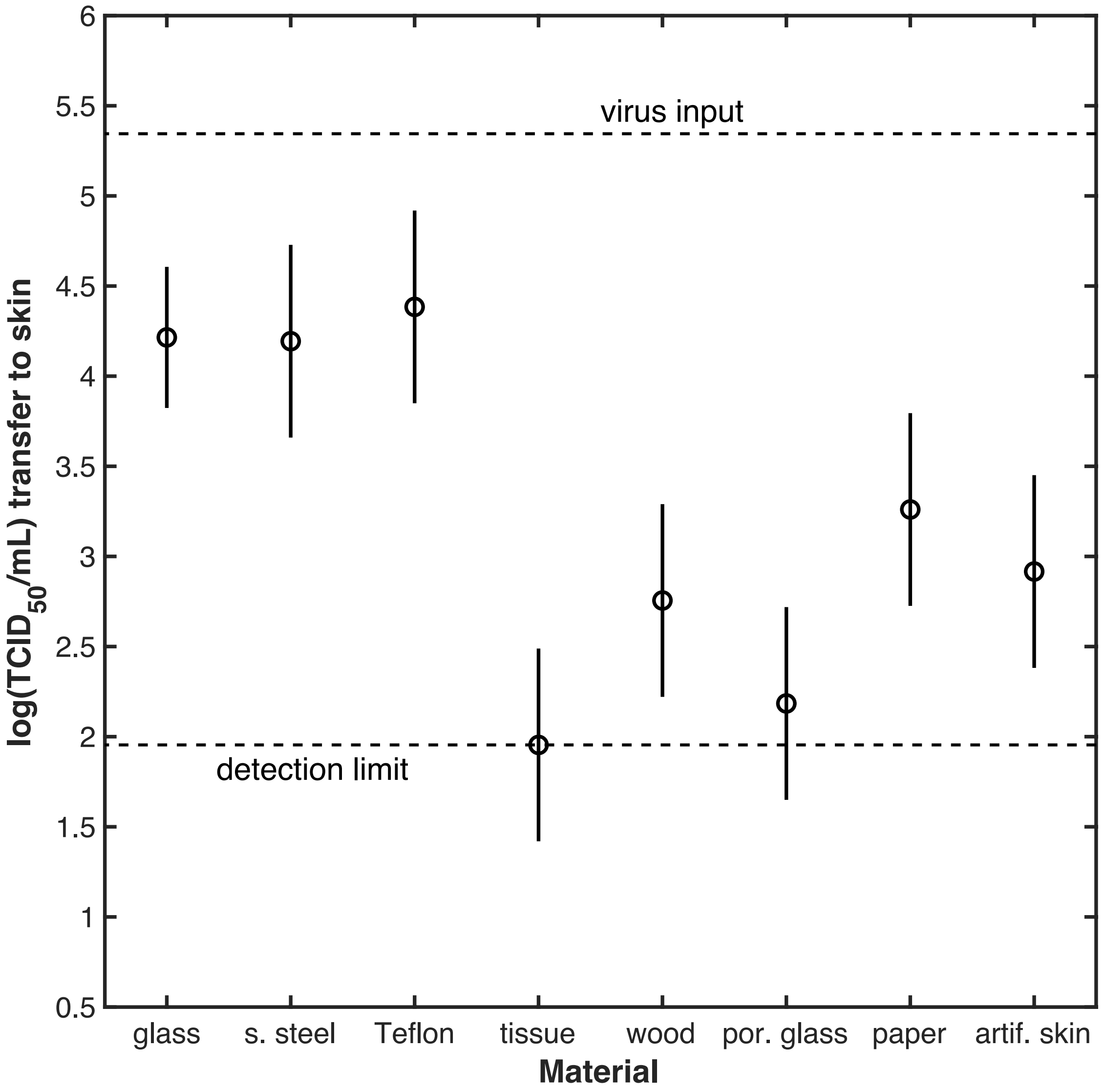

**Supplementary Figure 9. Post Hoc comparison of log($TCID_{50}$/mL) transferred to skin after contact with different solid samples.** The log($TCID_{50}$/mL) value includes results for both wet and dry conditions. The vertical bars represent comparison intervals; two groups are statistically different if their intervals do not overlap. Log($TCID_{50}$/mL) for each non-porous solid is greater than for each porous solids. We do not resolve a difference among the log($TCID_{50}$/mL) for the various impermeable solids.

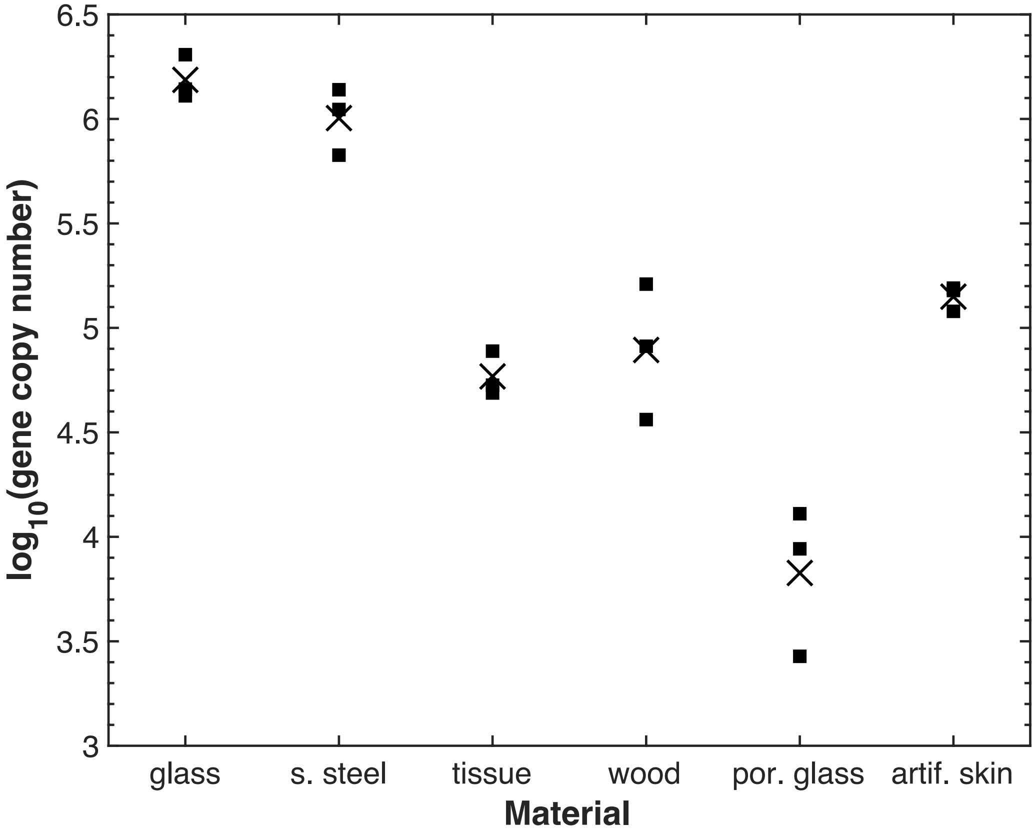

**Supplementary Figure 10. RT-qPCR results for RNA that was eluted by liquid extraction (See 5.4.6) from various solids 30 min. after a 1 µL droplet was placed on the solid.** Squares represent individual independent measurements and the X represent the mean of the log of three measurements. Supplementary Figure 11 explains the use of the log transformation. We know from prior work that steel and glass only slowly inactivate SARS-CoV-2[3], and the other materials do not have known active ingredients against the virus. We, therefore, interpret these results to be a test of the efficiency of liquid extraction of SARS-CoV-2. Comparing porous glass and glass, we see that it is more difficult to extract virus from a porous version of the same material. This is also generally true for other materials: recovery from all the porous solids is less than recovery of each of the non-porous solids. For the transfer experiments, “extraction” from the solid was done via solid-solid contact, which is distinct from liquid phase extraction, but the results here indicate that the virus enters the pores and therefore would not be available on the surface to be transferred.

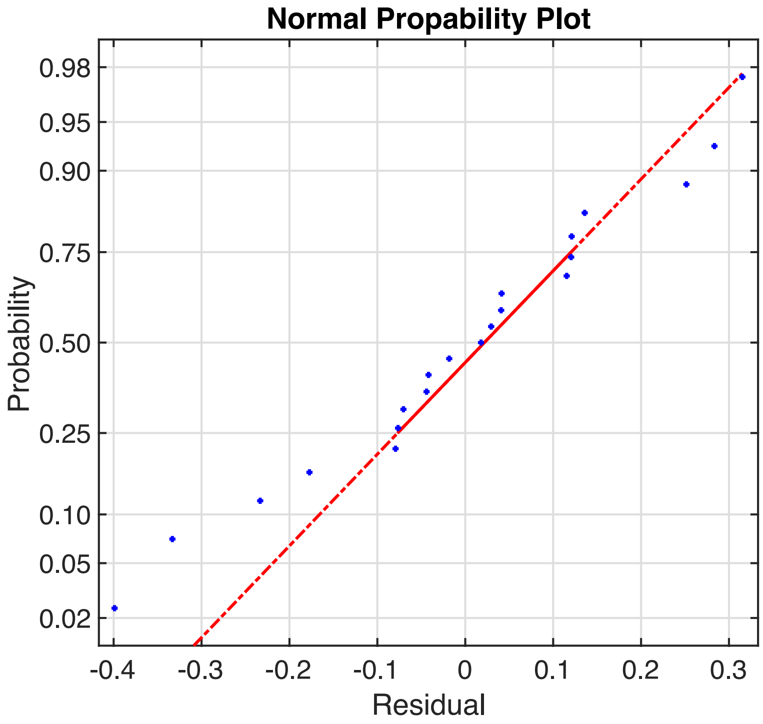

**Supplementary Figure 11. Normal probability plot (bottom) of residuals of qPCR data after a log10 transform.** The residuals are approximately normally distributed, although deviations at large magnitude of residual are noted.

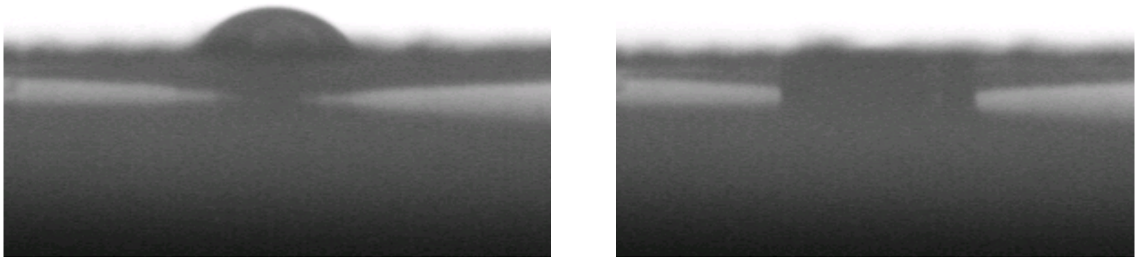

**Supplementary Figure 12. Image of water droplet on paper sample.** Freshly placed droplet (left) and after 2.5 minutes (right). The droplet initially sits on the surface, but after about 2.5 minutes it imbibes into the solid. The droplet volume is 1 µL.

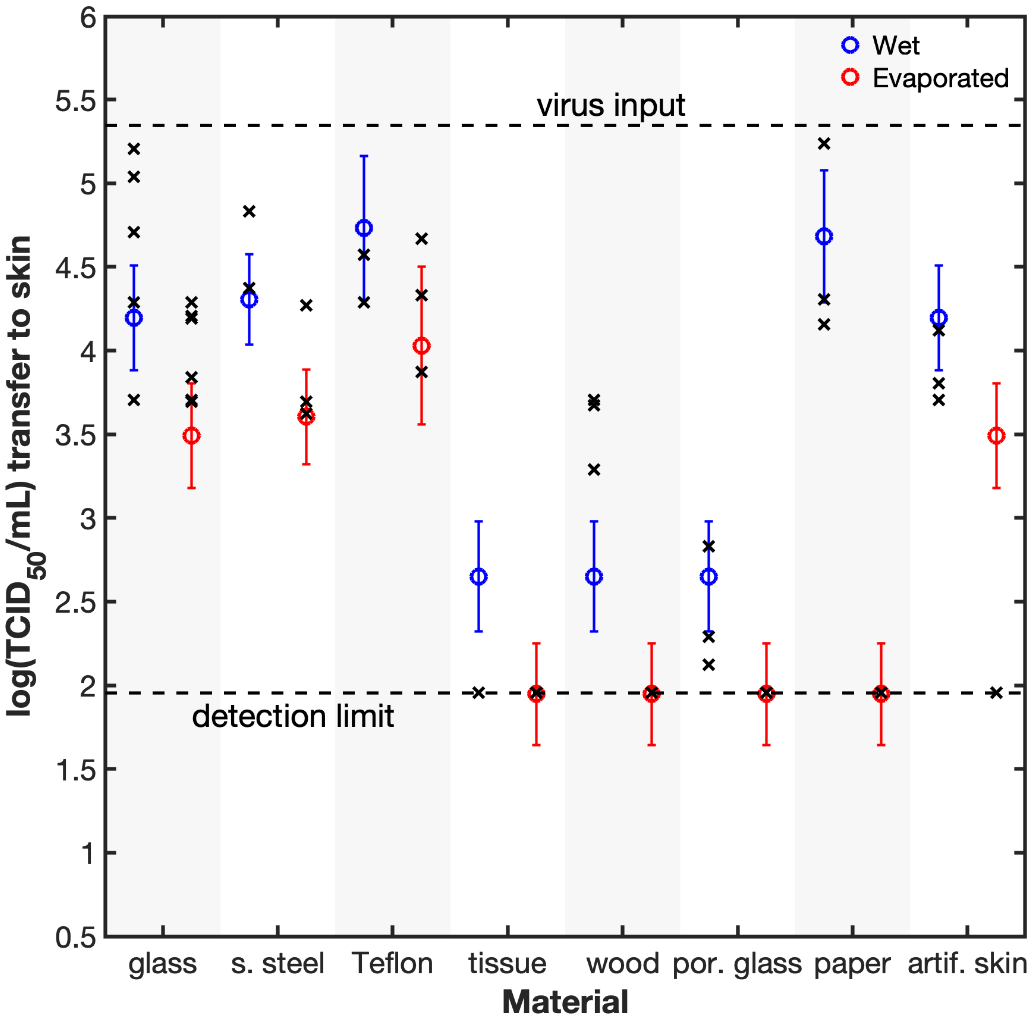

**Supplementary Figure 13. Predictive model of the transfer.** Linear regression using the programming language, R, was used to make a predictive model for the $TCID_{50}$/mL, with the following factors: $x$ = wet /dry (categorical), $y$ = cos(advancing contact angle), and $z$ = permeable/impermeable (categorical). Interaction terms were not found to be significant. The fitted equation was:

$$\text{Log } [TCID_{50}/mL] = 0.7025\, x - 0.4889\, y - 1.544\, z + 3.9808$$

And the p-values for the coefficients are 0.000113, 0.041, and $6.6 \times 10^{-11}$, respectively for wetness, contact angle and permeability, and $<10^{-6}$ for the intercept. The coefficient and p-value for contact angle were sensitive to our choice of the number to use for "undetectable". If we used 9.5 $TCID_{50}$/mL instead of 90, then the wettability coefficient became insignificant, so this coefficient may not be significant. Despite that, the model provides a good prediction of the $TCID_{50}$, as seen in the figure. One point of failure is the discrepancy for dry skin. Skin is rough so much of the virus in the dried state may be in crevices and therefore unavailable for transfer. If we were to include roughness as a factor, or broaden the concept of porosity to porosity or roughness, then clearly the model would perform better.
